# Plasma Taurine Relative Abundance, Not Dietary Intake or Genetic Predisposition, Predicts All-Cause Mortality and Unhealthy Ageing: A Prospective Cohort Study

**DOI:** 10.64898/2026.07.09.26357704

**Authors:** Jieun Lyu, Su-Jin Lee, Ji-Yun Hwang, Joong-Yeon Lim, Yoon Jung Park

## Abstract

**Background:** The influence of taurine on biological ageing remains unclear, particularly whether it acts as a causal driver or a functional biomarker. We aimed to disentangle the distinct roles of plasma taurine relative abundance, dietary taurine supply, and genetic metabolic capacity on all-cause mortality and unhealthy ageing.

**Methods:** This prospective study used data from the Korean Genome and Epidemiology Study (2001–2022). A subcohort of 2,321 participants (mean age 56.5 years; 51.4% female) with complete metabolomic, dietary, and genomic data was analyzed. Three independent pathways were evaluated: (1) plasma taurine/total amino acid (AA) ratio, (2) dietary taurine-to-protein ratio, and (3) a weighted genetic risk score (GRS) from 21 SNPs in taurine biosynthesis and transport genes. Primary outcomes were all-cause mortality and unhealthy ageing (Physiological Healthy Ageing Index [PHAI] score ≤25th percentile).

**Results:** A higher plasma taurine/total AA ratio was consistently associated with improved ageing outcomes. Participants in the highest quartile showed 29% lower all-cause mortality (Hazard Ratio [HR], 0.71; 95% Confidence Interval [CI], 0.52–0.98; *P* for trend = .04) and lower risk of PHAI-based unhealthy ageing (HR, 0.77; 95% CI, 0.59–1.00; *P* for trend = .04) versus the lowest quartile. Dietary taurine-to-protein ratio was not associated with mortality (*P* for trend = .70), nor was the GRS (*P* for trend = .74).

**Conclusions:** The protective association of taurine was linked to its relative abundance within the systemic amino acid pool, rather than dietary intake or genetic predisposition, supporting taurine as a functional biomarker of metabolic efficiency rather than a deterministic causal driver of ageing.

**Key Points:**

- Higher plasma taurine relative abundance was associated with lower risks of all-cause mortality and PHAI-based unhealthy ageing.
- Dietary taurine intake relative to protein consumption was not associated with unhealthy ageing outcomes.
- Genetic predisposition for taurine metabolism showed no significant association with mortality or unhealthy ageing.
- Preservation of taurine within the systemic amino acid pool may be more important than dietary intake or endogenous synthesis.

## Introduction

Taurine (2-aminoethanesulfonic acid), the most abundant free amino acid in mammalian tissues, fulfils diverse physiological functions, including osmoregulation, membrane stabilization, intracellular calcium modulation, bile acid conjugation, and mitochondrial transfer ribonucleic acid modification [1–7]. Circulating taurine levels are governed by three independent determinants: exogenous supply from animal-derived foods (meat and seafood) [8, 9]; endogenous biosynthesis via the transsulfuration pathway (methionine → cysteine → taurine), catalysed by cysteine dioxygenase 1 (*CDO1*) and cysteine sulfinic acid decarboxylase (*CSAD*) [10, 11]; and renal reabsorption mediated by the sodium- and chloride-dependent transporter *SLC6A6* [12]. This tripartite regulation means that circulating taurine reflects the combined effects of dietary intake, biosynthetic capacity, and transport efficiency, rather than any single biological process. This complexity has been largely overlooked in studies investigating the role of taurine in human ageing.

Epidemiological evidence consistently links lower plasma taurine to a wide range of age-related chronic diseases, [13–17] and taurine supplementation has been shown to reduce key components of metabolic syndrome in randomised controlled trials [18]. These findings received prominent mechanistic support from Singh et al. [19], who demonstrated that taurine supplementation extended both health span and lifespan in mice and improved health span in non-human primates, suggesting that taurine deficiency may be a causal driver of ageing. However, this interpretation was directly challenged by Fernandez et al. [20], who used longitudinal data from three healthy human cohorts, as well as non-human primates and mice, to demonstrate that circulating taurine concentrations increased or remained unchanged with age, thereby undermining the premise that taurine deficiency accumulates with ageing in humans.

Nevertheless, an important gap remains in the existing literature: to our knowledge, no previous studies have systematically examined the independent contributions of dietary taurine supply, genetic metabolic capacity, and endogenous taurine homeostasis to ageing outcomes within a single analytical framework. Thus, addressing this gap may help clarify whether taurine functions as a driver or a marker of ageing in humans.

To address this gap, we used data from the Korean Genome and Epidemiology Study (KoGES) community-based cohort, which integrates plasma metabolomics, genome-wide genotyping, and repeated dietary assessments, to evaluate the independent contributions of three taurine-related exposures to all-cause mortality and PHAI-based unhealthy ageing: (1) plasma taurine relative abundance (taurine/total amino acid [AA] ratio), (2) dietary taurine-to-protein ratio, and (3) a weighted genetic risk score (GRS) derived from single nucleotide polymorphisms (SNPs) in taurine-specific genes.

## Methods

### Study Population

The KoGES community-based cohort is a prospective population-based study initiated in 2001 that included adults aged 40–69 years residing in urban (Ansan) and rural (Anseong) communities in Gyeonggi, South Korea [21]. A total of 10,030 individuals were enrolled at baseline (2001–2002), with biennial follow-up until 2019–2020 (up to 14 years).

In this study, a subsample of 2,580 participants was selected as a metabolomics subcohort. After excluding participants with missing Physiology Healthy Ageing Index (PHAI) components (n = 220), implausible energy intake (n = 16), and missing covariates (n = 23), 2,321 were included in the primary analysis. The GRS analysis included 2,144 participants with available genotyping data passing quality control. For PHAI analyses, individuals with an unhealthy ageing status at baseline were excluded, yielding 1,185 metabolomic/dietary and 1,135 GRS participants for healthy ageing analyses (Figure 1). The study was approved by an Institutional Review Board (approval numbers: 2018-03-05-4C-A; 2022-07-07-C-A; ewha-202510-0042-01) and conducted in accordance with the Declaration of Helsinki. Written informed consent was obtained from all participants.

**Figure 1.**
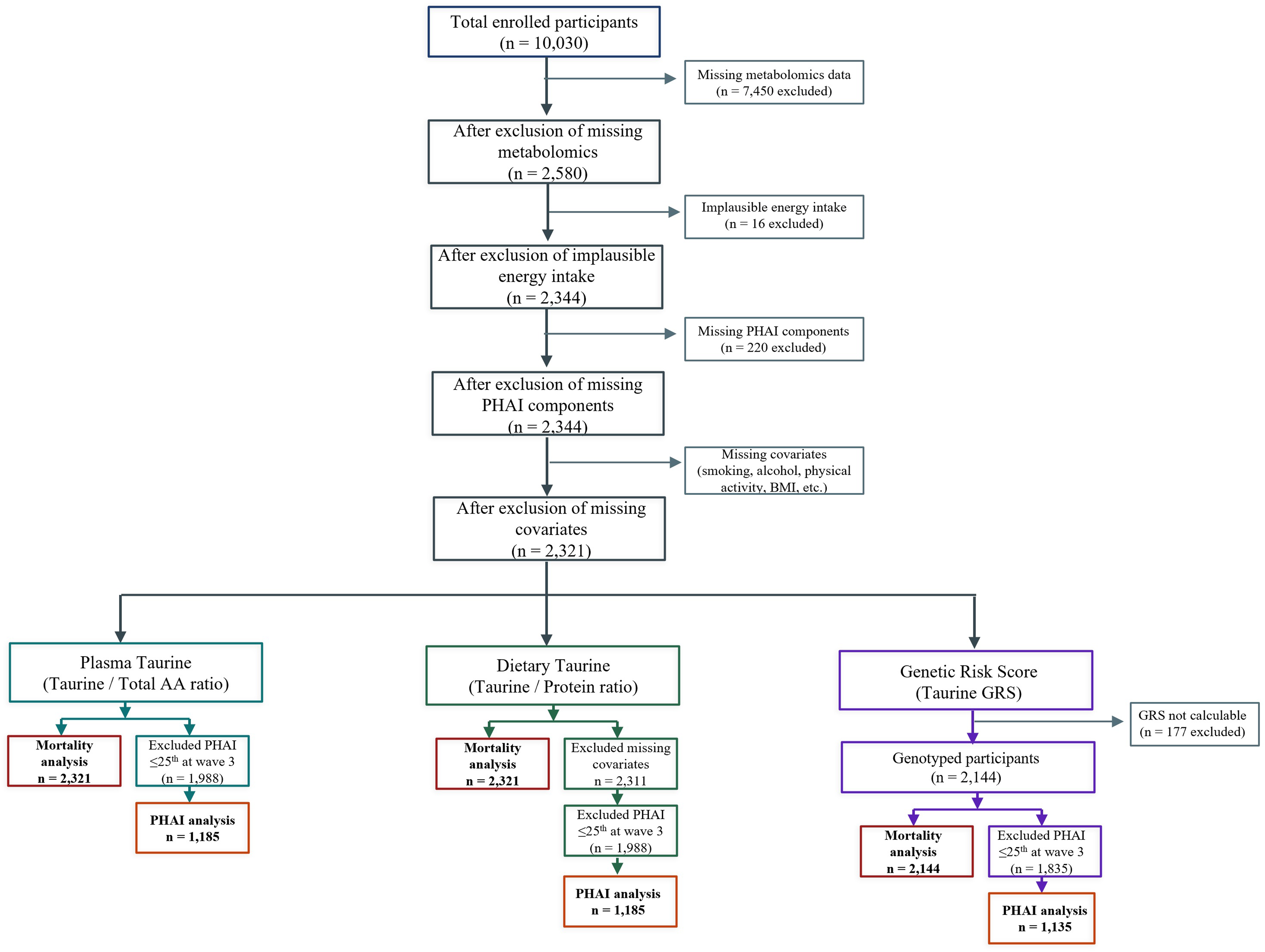
Study design and participant flow diagram. Flowchart showing the three parallel analytical cohorts derived from the Korean Genome and Epidemiology Study community-based cohort: the metabolomics sub cohort (n = 2,321 for primary analyses), the dietary analysis cohort (n = 2,564), and the genetic analysis cohort (n = 2,144). **Abbreviations:** BMI, body mass index; GRS, genetic risk score; PHAI, Physiology Healthy Ageing Index; taurine/total AA ratio, plasma taurine relative abundance.

### Exposure Variables

Plasma metabolites were quantified using the AbsoluteIDQ p180 kit (Biocrates Life Sciences AG, Innsbruck, Austria), [22] measuring 22 amino acids and related metabolites via flow injection analysis- and liquid chromatography-tandem mass spectrometry (LC-MS/MS). The primary plasma exposure, taurine/total AA ratio, was calculated as plasma taurine divided by the sum of 22 circulating amino acids, providing a normalised index of taurine homeostasis within the total amino acid pool that accounts for overall nutritional status and amino acid pool size. As a sensitivity analysis, a taurine/methionine ratio was calculated using methionine as the upstream transsulfuration pathway precursor (Supplementary Tables S4–S6).

Dietary intake was estimated by linking a validated 106-item food frequency questionnaire (FFQ) [23] with a comprehensive database developed through sequential linkage with the Korea National Health and Nutrition Examination Survey (2022–2023) and the Korean Food Composition Tables. Each FFQ item was disaggregated into 478 components to incorporate amino acid profiles not originally available in the cohort [24]. The taurine-to-protein ratio was used to account for its exclusive presence in animal-derived proteins.

The weighted GRS was constructed from 21 SNPs in *CDO1* (5q23.2), *CSAD* (12q13.13), and *SLC6A6* (3p24.2) passing quality control (genotype call rate ≥ 95%; minor allele frequency ≥ 0.01; Hardy–Weinberg equilibrium *P* > 1×10DD) [25–27]. SNP weights (beta coefficients) were estimated from multivariable linear regression of plasma taurine adjusted for age, sex, and dietary taurine intake. A GRS × dietary taurine intake interaction score was calculated to examine effect modification by dietary taurine supply. A sensitivity 117-SNP GRS incorporating upstream pathway genes (*CBS* and *CTH*) and an accessory transporter (*SLC36A2*) [28–32] was constructed for sensitivity analyses. All exposures were categorised into quartiles (Q1–Q4).

### Outcomes

Survival status was determined through probabilistic record linkage with the Korean National Death Registry. Causes of death were classified by the Korean Standard Classification of Diseases, Sixth Revision (based on the International Classification of Diseases, Tenth Revision [ICD-10]) [33, 34]. Mortality outcomes included all-cause mortality, age-related mortality (excluding infectious diseases, meningitis, encephalitis, and wounds), cancer mortality (ICD-10: C00–C97), and cardiovascular disease (CVD) mortality (ICD-10: I00–I99).

Biological ageing was assessed using the PHAI, [35] a validated composite index integrating five physiological parameters: systolic blood pressure, fasting blood glucose, serum creatinine, forced vital capacity, and c-reactive protein (CRP)/high-sensitivity CRP, each scored from 0–2 based on established clinical cutoffs, yielding a total score between 0 and 10 (lower scores = poorer physiological health). PHAI-based unhealthy ageing was defined as a PHAI score ≤ 25th percentile of the baseline distribution occurring during follow-up.

### Statistical Analysis

Cox proportional hazards models estimated hazard ratios (HRs) and 95% confidence intervals (CIs) for Q2–Q4 versus Q1. Tests for linear trend were assigned quartile median values as a continuous variable. Models were unadjusted (crude), adjusted for age and sex (Model 1), then further adjusted for education, household income, smoking, alcohol use, physical activity, body mass index (BMI), total energy intake, and dietary taurine intake or protein intake, as appropriate (Model 2, fully adjusted). Sex was determined based on self-reported data obtained at enrolment and was treated as a biological variable in all analyses, with sex-stratified analyses additionally adjusted for menopausal status in females. The proportional hazards assumption was assessed using Schoenfeld residuals.

All analyses were performed using SAS (version 9.4; SAS Institute Inc, Cary, NC, USA), with a two-sided *P* < .05 indicating statistical significance. This study was reported in accordance with the Strengthening the Reporting of OBservational studies in Epidemiology (STROBE) guidelines for observational research.

## Results

### Study Participants

The mean age of participants was 56.5 years (range 43–74 years); 51.4% were female. Table 1 presents the baseline characteristics stratified by plasma taurine/total AA quartiles. Q4 participants were more likely to be female and older compared with participants in Q1. PHAI scores showed a stepwise decrease across quartiles, suggesting that higher taurine relative abundance was associated with somewhat lower physiological health status at baseline. Q4 participants also had higher waist circumference, systolic blood pressure, and total cholesterol than participants in Q1; however, all these measurements were within clinically normal ranges. After energy adjustment, no significant between-group differences were observed for dietary taurine or amino acid intakes across quartiles (all *P*-trend > .05), indicating that differences in total energy intake rather than selective taurine intake explained the dietary variation.

**Table 1.**
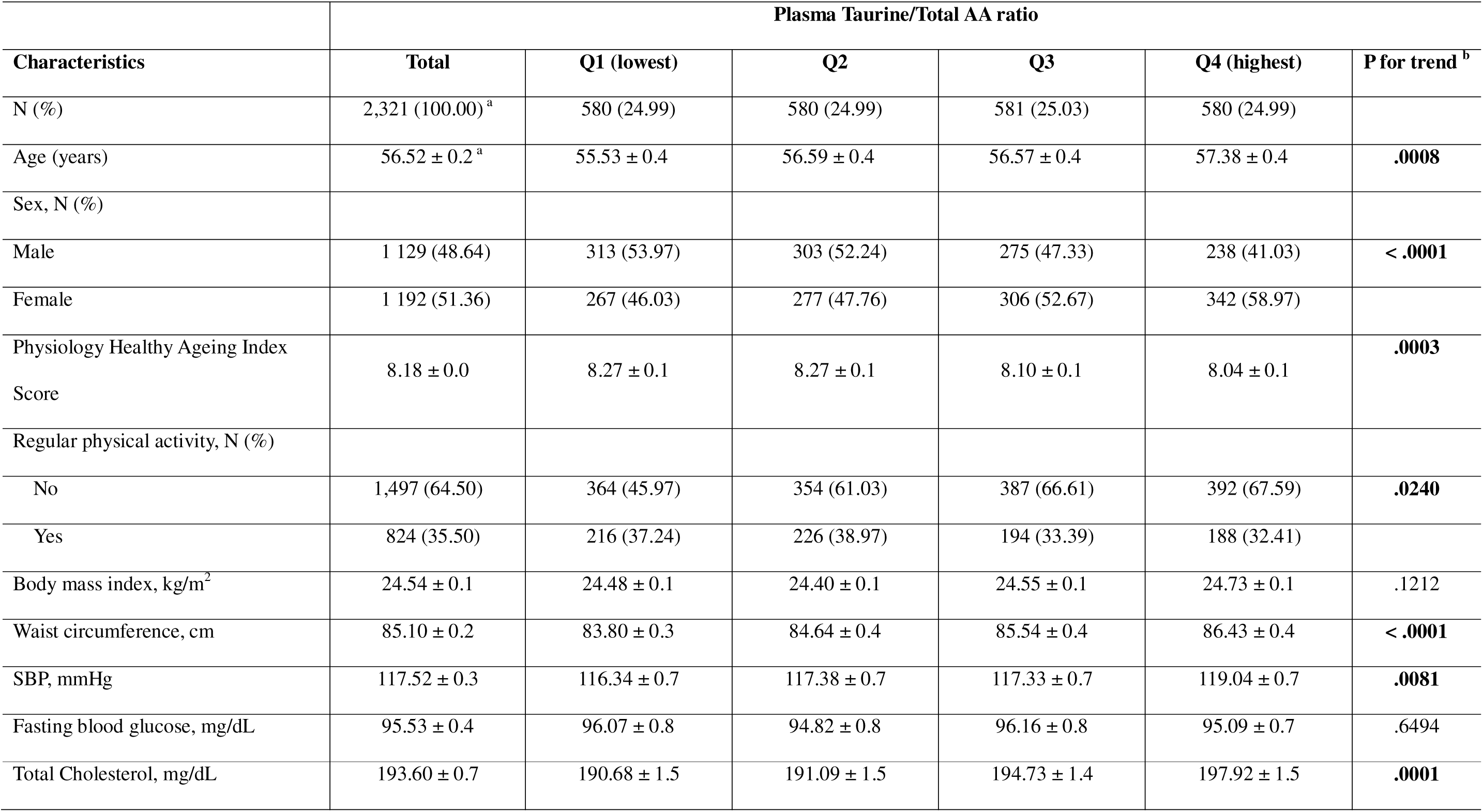
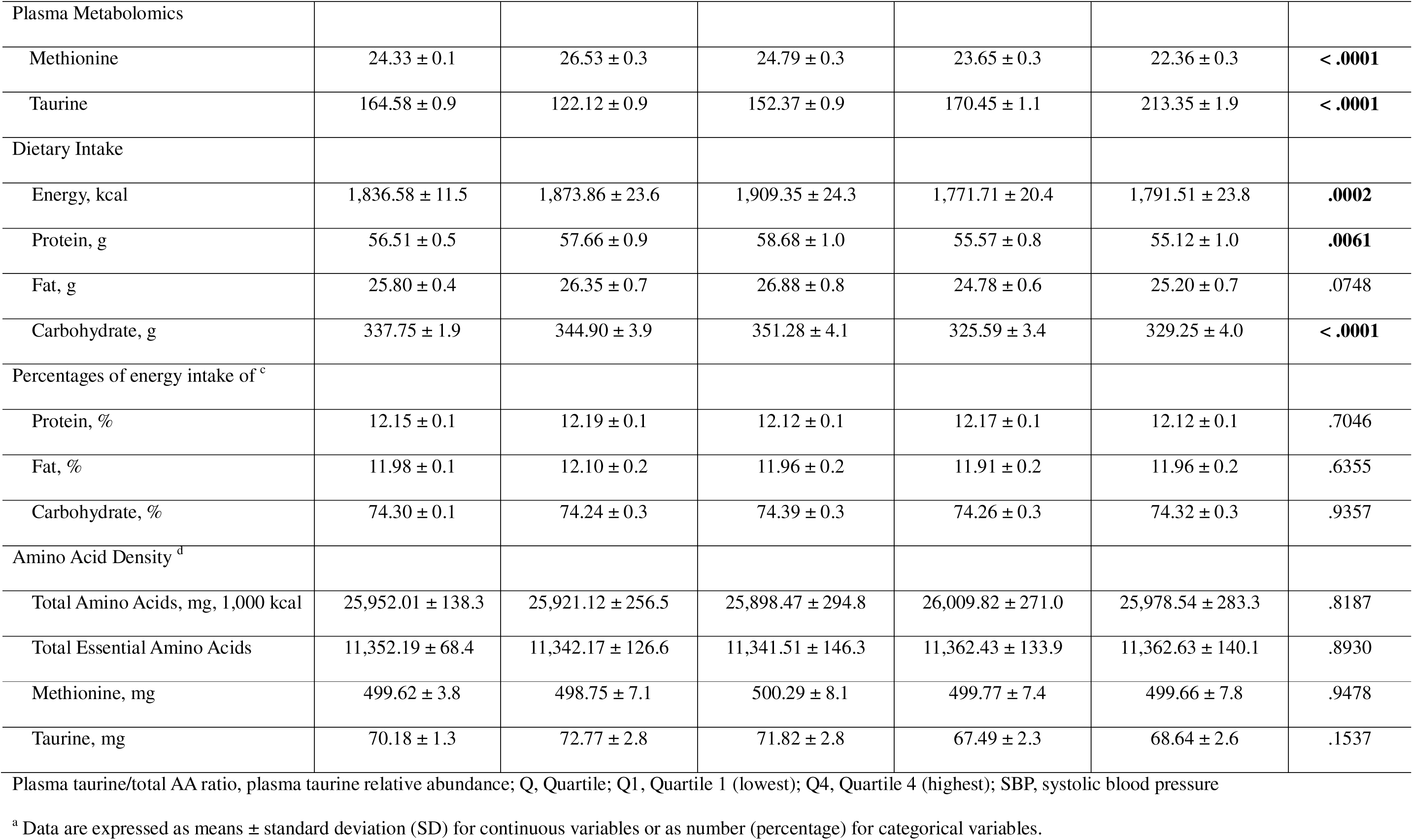
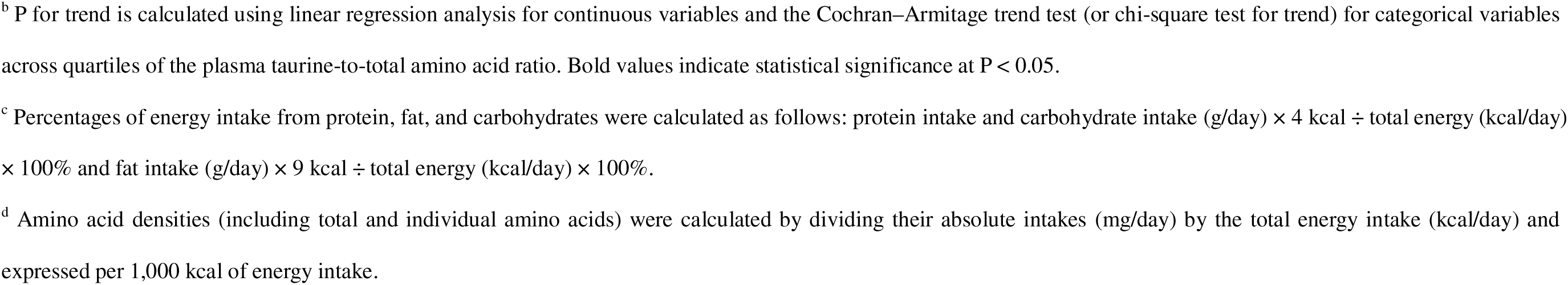
Baseline characteristics of study participants stratified by quartiles of plasma taurine relative abundance (n = 2,321).

### Plasma Taurine/Total AA Ratio and Ageing Outcomes

A higher plasma taurine/total AA ratio was consistently associated with reduced ageing-related outcomes in the fully adjusted model (Table 2). Compared with Q1, Q4 showed a 29% lower all-cause mortality risk (HR, 0.71, 95% CI 0.52–0.98; *P*-trend = .038) and similar reductions for age-related mortality (HR, 0.68; *P*-trend = .037) and cancer mortality (HR, 0.54, 95% CI 0.33–0.90; *P*-trend = .025). No significant association was observed for CVD mortality. Importantly, these associations were independent of dietary taurine intake, indicating that the observed benefit reflects endogenous metabolic status. For PHAI-based unhealthy ageing, Q4 showed a 23% reduction in risk (HR, 0.77, 95% CI 0.59–1.00; *P*-trend = .041), despite Q4 participants exhibiting somewhat less favourable baseline clinical profiles than participants in the other quartiles, a pattern consistent with physiological resilience (Table 3).

**Table 2.** Multivariable-adjusted hazard ratios (HRs) and 95% confidence intervals (CIs) for all-cause and cause-specific mortality according to quartiles of plasma taurine-to-total amino acid ratio.

|  | Plasma Taurine/Total AA ratio |  |  |  |  |
| --- | --- | --- | --- | --- | --- |
| HRs (95% CIs) | Q1 (lowest) | Q2 | Q3 | Q4 (highest) | P for trend |
| No. of at risk | 580 | 580 | 581 | 580 |  |
| Person-years | 7,946 | 7,809 | 8,093 | 7,994 |  |
| Incidence rate per 1000 person-years | 10.45 | 9.35 | 9.89 | 9.76 |  |
| All-cause |  |  |  |  |  |
| No. of event | 83 | 73 | 80 | 78 |  |
| Crude model | Reference | 0.94 (0.69–1.29) | 0.93 (0.68–1.26) | 0.94 (0.69–1.28) | .7114 |
| Model 1 | Reference | 0.85 (0.62–1.16) | 0.83 (0.61–1.13) | 0.79 (0.58–1.08) | .1500 |
| Model 2 | Reference | 0.84 (0.61–1.16) | 0.80 (0.58–1.09) | 0.71 (0.52–0.98) | **.0376** |
| Age-related |  |  |  |  |  |
| No. of event | 72 | 60 | 68 | 65 |  |
| Crude model | Reference | 0.88 (0.63–1.24) | 0.91 (0.65–1.27) | 0.90 (0.65–1.26) | .5989 |
| Model 1 | Reference | 0.79 (0.56–1.11) | 0.82 (0.59–1.14) | 0.75 (0.54–1.06) | .1275 |
| Model 2 | Reference | 0.79 (0.56–1.12) | 0.79 (0.56–1.11) | 0.68 (0.49–0.96) | **.0371** |
| Cancer |  |  |  |  |  |
| No. of event | 38 | 25 | 29 | 26 |  |
| Crude model | Reference | 0.68 (0.41–1.13) | 0.74 (0.46–1.21) | 0.68 (0.41–1.12) | .1553 |
| Model 1 | Reference | 0.62 (0.38–1.03) | 0.70 (0.43–1.13) | 0.60 (0.37–1.00) | .0653 |
| Model 2 | Reference | 0.60 (0.36–1.00) | 0.66 (0.40–1.07) | 0.54 (0.33–0.90) | **.0252** |
| CVD |  |  |  |  |  |
| No. of event | 16 | 15 | 18 | 22 |  |
| Crude model | Reference | 1.01 (0.50–2.04) | 1.08 (0.55–2.11) | 1.38 (0.73–2.63) | .2874 |
| Model 1 | Reference | 0.89 (0.44–1.80) | 0.93 (0.47–1.84) | 1.08 (0.57–2.07) | .7250 |
| Model 2 | Reference | 0.94 (0.46–1.93) | 0.97 (0.49–1.94) | 1.00 (0.51–1.95) | .9679 |

plasma taurine/total AA ratio, plasma taurine relative abundance; Q, Quartile; Q1, Quartile 1 (lowest); Q4, Quartile 4 (highest); HR, hazard ratio; CI, confidence interval; CVD, cardiovascular disease.
<sup>a</sup> Crude model was unadjusted; Model 1 was adjusted for age (years, continuous) and sex; Model 2 was further adjusted for education level (elementary school or below, middle school, high school, or college or above), household income level (quartiles), smoking status (never, former, or current smoker), drinking status (never, former, or current drinker), regular physical activity (yes or no), body mass index (kg/m<sup>2</sup>, continuous), total energy intake (kcal/day, continuous), and dietary taurine intake (mg/day, continuous). <sup>b</sup> P for trend was calculated by assigning the median value of each quartile to participants as a continuous variable across all models. Bold values indicate statistical significance at P < 0.05.

**Table 3.**
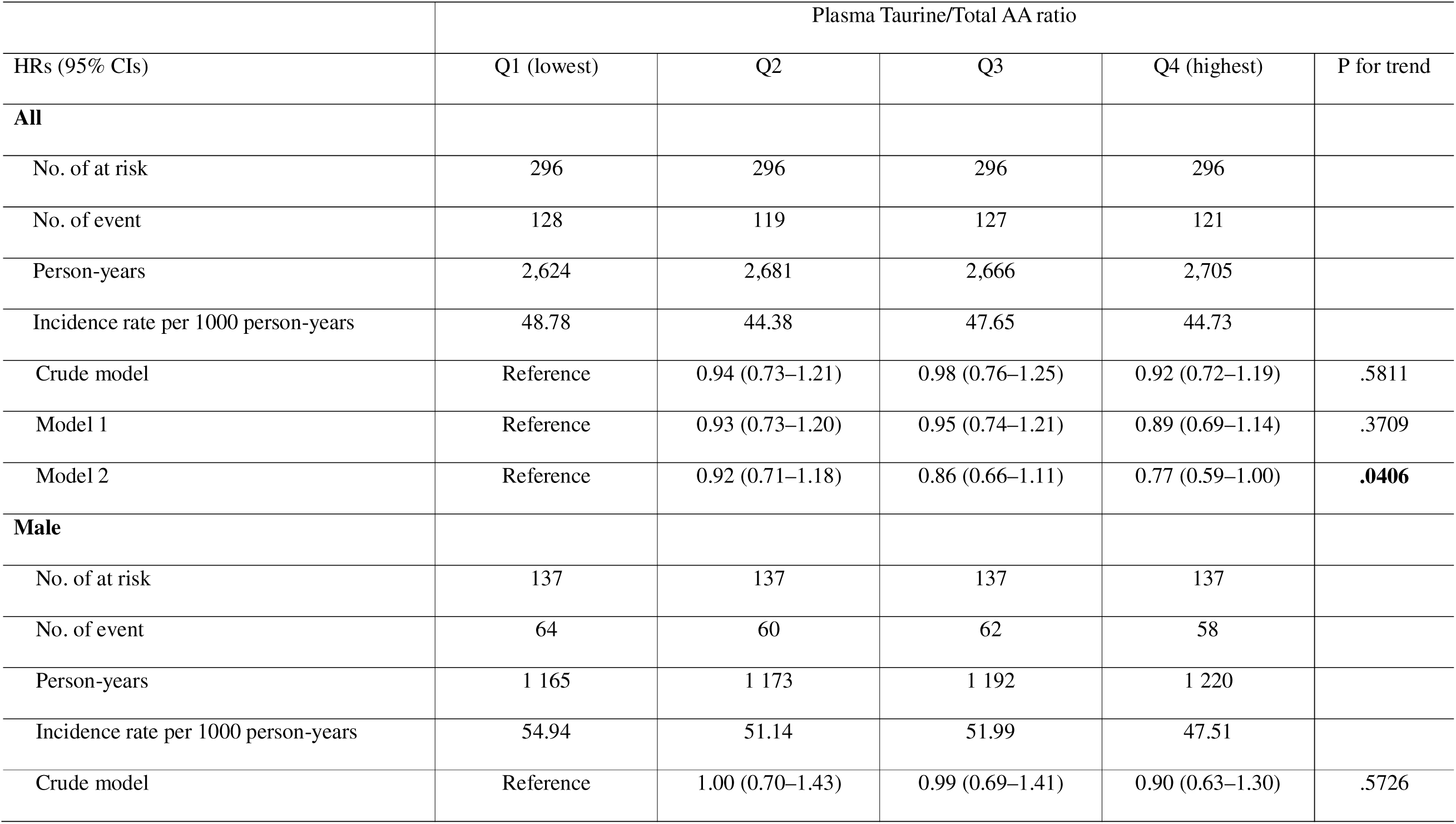
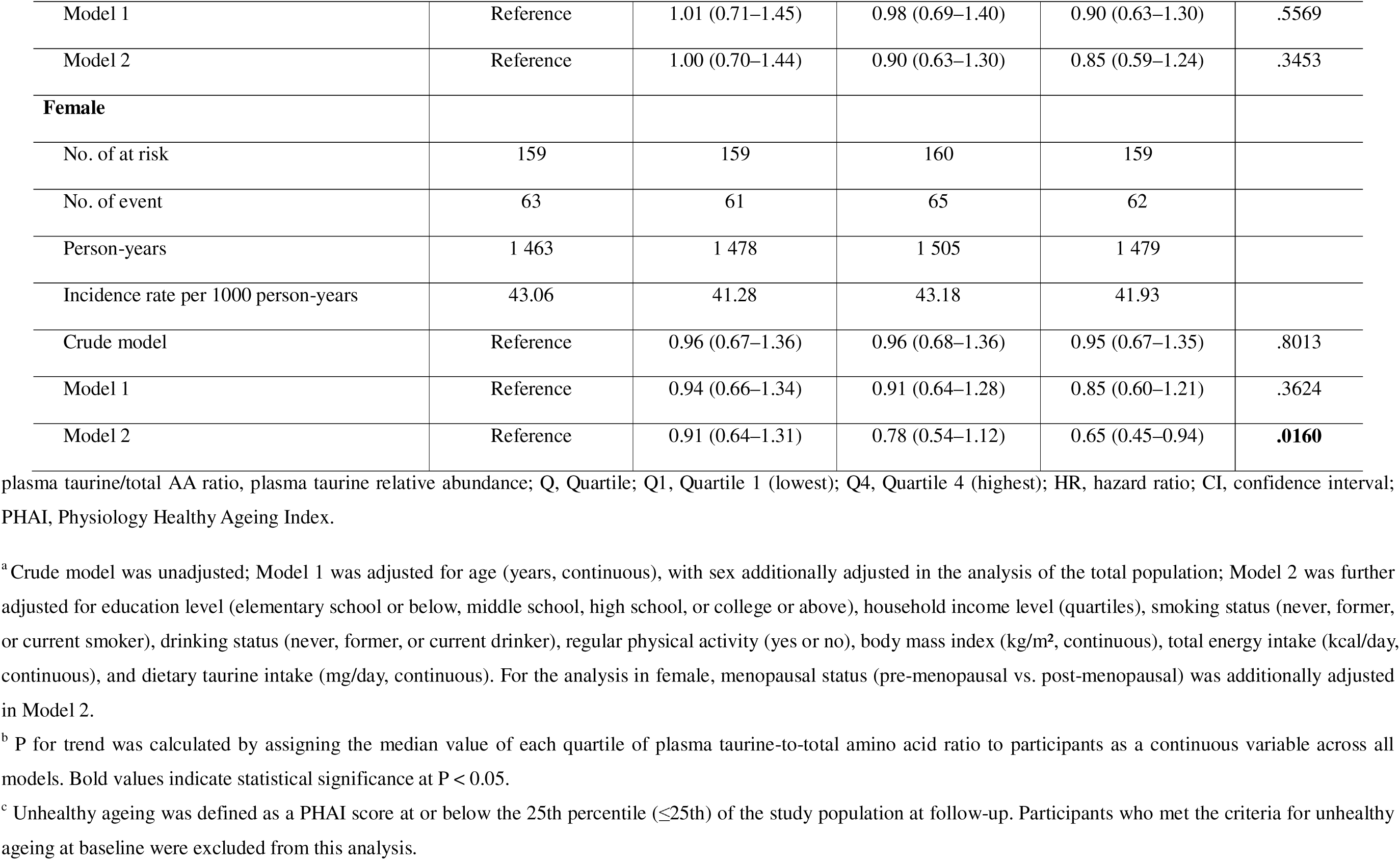
Multivariable-adjusted hazard ratios (HRs) and 95% confidence intervals (CIs) for the incidence of PHAI-based unhealthy ageing according to quartiles of plasma taurine-to-total amino acid ratio.

|  | Plasma Taurine/Total AA ratio |  |  |  |  |
| --- | --- | --- | --- | --- | --- |
| HRs (95% CIs) | Q1 (lowest) | Q2 | Q3 | Q4 (highest) | P for trend |
| **All** |  |  |  |  |  |
| No. of at risk | 296 | 296 | 296 | 296 |  |
| No. of event | 128 | 119 | 127 | 121 |  |
| Person-years | 2,624 | 2,681 | 2,666 | 2,705 |  |
| Incidence rate per 1000 person-years | 48.78 | 44.38 | 47.65 | 44.73 |  |
| Crude model | Reference | 0.94 (0.73–1.21) | 0.98 (0.76–1.25) | 0.92 (0.72–1.19) | .5811 |
| Model 1 | Reference | 0.93 (0.73–1.20) | 0.95 (0.74–1.21) | 0.89 (0.69–1.14) | .3709 |
| Model 2 | Reference | 0.92 (0.71–1.18) | 0.86 (0.66–1.11) | 0.77 (0.59–1.00) | **.0406** |
| **Male** |  |  |  |  |  |
| No. of at risk | 137 | 137 | 137 | 137 |  |
| No. of event | 64 | 60 | 62 | 58 |  |
| Person-years | 1 165 | 1 173 | 1 192 | 1 220 |  |
| Incidence rate per 1000 person-years | 54.94 | 51.14 | 51.99 | 47.51 |  |
| Crude model | Reference | 1.00 (0.70–1.43) | 0.99 (0.69–1.41) | 0.90 (0.63–1.30) | .5726 |
| Model 1 | Reference | 1.01 (0.71–1.45) | 0.98 (0.69–1.40) | 0.90 (0.63–1.30) | .5569 |
| Model 2 | Reference | 1.00 (0.70–1.44) | 0.90 (0.63–1.30) | 0.85 (0.59–1.24) | .3453 |
| **Female** |  |  |  |  |  |
| No. of at risk | 159 | 159 | 160 | 159 |  |
| No. of event | 63 | 61 | 65 | 62 |  |
| Person-years | 1 463 | 1 478 | 1 505 | 1 479 |  |
| Incidence rate per 1000 person-years | 43.06 | 41.28 | 43.18 | 41.93 |  |
| Crude model | Reference | 0.96 (0.67–1.36) | 0.96 (0.68–1.36) | 0.95 (0.67–1.35) | .8013 |
| Model 1 | Reference | 0.94 (0.66–1.34) | 0.91 (0.64–1.28) | 0.85 (0.60–1.21) | .3624 |
| Model 2 | Reference | 0.91 (0.64–1.31) | 0.78 (0.54–1.12) | 0.65 (0.45–0.94) | **.0160** |

plasma taurine/total AA ratio, plasma taurine relative abundance; Q, Quartile; Q1, Quartile 1 (lowest); Q4, Quartile 4 (highest); HR, hazard ratio; CI, confidence interval; PHAI, Physiology Healthy Ageing Index.
<sup>a</sup> Crude model was unadjusted; Model 1 was adjusted for age (years, continuous), with sex additionally adjusted in the analysis of the total population; Model 2 was further adjusted for education level (elementary school or below, middle school, high school, or college or above), household income level (quartiles), smoking status (never, former, or current smoker), drinking status (never, former, or current drinker), regular physical activity (yes or no), body mass index (kg/m<sup>2</sup>, continuous), total energy intake (kcal/day, continuous), and dietary taurine intake (mg/day, continuous). For the analysis in female, menopausal status (pre-menopausal vs. post-menopausal) was additionally adjusted in Model 2.
<sup>b</sup> P for trend was calculated by assigning the median value of each quartile of plasma taurine-to-total amino acid ratio to participants as a continuous variable across all models. Bold values indicate statistical significance at $P < 0.05$ .
<sup>c</sup> Unhealthy ageing was defined as a PHAI score at or below the 25th percentile ( $\leq 25$ th) of the study population at follow-up. Participants who met the criteria for unhealthy ageing at baseline were excluded from this analysis.

Sex-stratified analyses revealed divergent patterns. Among males, a higher taurine/total AA ratio was significantly associated with reduced all-cause mortality (HR, 0.67; *P*-trend = .027), with a consistent but non-significant trend for PHAI-based unhealthy ageing (Supplementary Tables S1 and S3). Among females, the protective association was statistically significant for PHAI-based unhealthy ageing (HR, 0.65, 95% CI 0.45–0.94; *P*-trend = .016), while no significant association was observed for mortality (Supplementary Tables S1 and S3). This sex-specific pattern (survival benefit among males and healthy ageing benefit among females) suggests that taurine homeostasis intersects with sex-specific ageing biology.

### Dietary Taurine-to-Protein Ratio and Ageing Outcomes

A higher dietary taurine-to-protein ratio was not significantly associated with all-cause, age-related, or CVD mortality in the total population or by sex after full adjustment (Figure 2; Supplementary Table S2). In sex-stratified analyses, males in Q4 showed a significantly reduced cancer mortality risk (HR, 0.48, 95% CI 0.23–1.02; *P*-trend = .047). Paradoxically, in the total population, Q4 of the dietary taurine-to-protein ratio was associated with a significantly increased risk of PHAI-based unhealthy ageing (HR, 1.32, 95% CI 1.02–1.71; *P*-trend = .041). This adverse association was markedly amplified in females (HR, 1.90, 95% CI 1.32–2.74; *P*-trend < .001), with no significant association in males. Critically, plasma taurine levels did not differ significantly across dietary quartiles, confirming that higher dietary intake did not translate into elevated circulating taurine.

**Figure 2.**
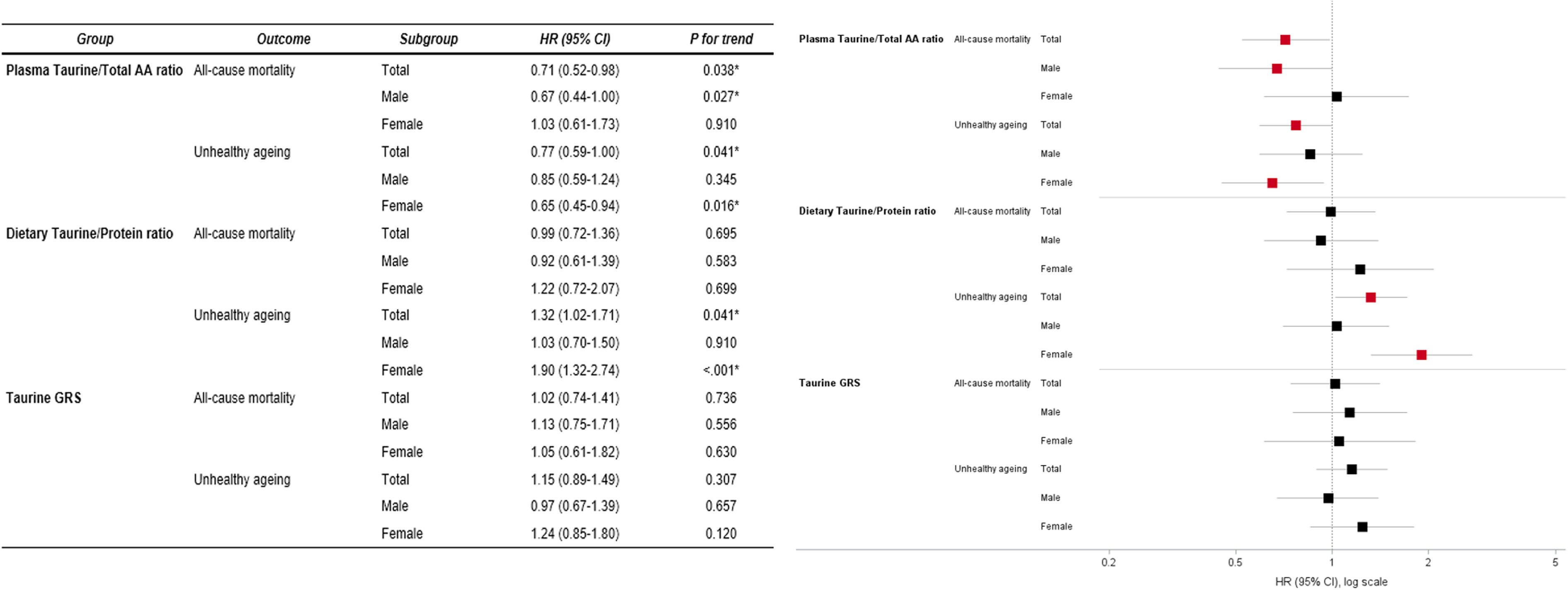
Associations of taurine pathways with all-cause mortality and PHAI-based unhealthy ageing. Forest plot summarizing the associations of the three taurine pathways (plasma taurine relative abundance, dietary taurine/protein ratio, and taurine genetic risk score; all Q4 vs. Q1) with all-cause mortality and unhealthy ageing (Physiology Healthy Ageing Index ≤ 25th percentile) in the total population and sex-stratified analyses. Squares represent hazard ratios, and horizontal lines represent 95% confidence intervals. The x-axis is presented on a log scale. Red squares indicate statistically significant associations (*P* for trend < .05), while black squares indicate non-significant associations. All models are adjusted for age, education level, household income, smoking status, drinking status, regular physical activity, body mass index, total energy intake, and dietary taurine intake. Models for females are additionally adjusted for menopausal status. *P* for trend is calculated by assigning the median value of each quartile as a continuous variable. **Abbreviations:** CI, confidence interval; GRS, genetic risk score; HR, hazard ratio; taurine/total AA ratio, plasma taurine relative abundance.

### Taurine GRS and Ageing Outcomes

The taurine GRS showed a stepwise increase in plasma taurine levels across quartiles (*P*-trend = .001), supporting its validity as a proxy for genetic taurine metabolic capacity (data not shown). However, the GRS was not significantly associated with all-cause mortality (*P*-trend = .736), any cause-specific mortality, or PHAI-based unhealthy ageing (*P*-trend = .307) in the total population or by sex (Figure 2; Supplementary Table S3). These null findings were consistent across both the primary 21-SNP GRS and the extended 117-SNP sensitivity GRS incorporating *CBS*, *CTH*, and *SLC36A2* (Supplementary Tables S7–S9). For the GRS × dietary taurine interaction analysis, a statistically significant adverse association was observed in females (Q4 interaction score: HR, 1.58, 95% CI 1.06–2.35; *P*-trend = .031), while no significant association was observed in males (*P*-trend = .838). This finding suggests that in females, the combination of high genetic taurine metabolic capacity and high dietary taurine intake may paradoxically increase PHAI-based unhealthy ageing risk.

## Discussion

To our knowledge, this study presents the first systematic triangulation of metabolomic, dietary, and genetic evidence regarding the role of taurine in human ageing within a large prospective cohort. Our findings demonstrated a coherent and internally consistent pattern: neither the dietary taurine-to-protein ratio nor genetic taurine metabolic capacity independently predicted all-cause mortality or PHAI-based unhealthy ageing. In contrast, the taurine/total AA ratio within the global amino acid pool was consistently and independently associated with reduced mortality and PHAI-based unhealthy ageing risk, even after adjustment for dietary taurine intake.

These findings directly address the unresolved controversy between Singh et al. [19] and Fernandez et al. [20]. The absence of a significant association between dietary taurine and ageing outcomes is consistent with the findings of Fernandez et al. [20], who concluded that external taurine supply does not reliably alter ageing trajectories in free-living populations. This discrepancy may reflect the fundamental differences between controlled high-dose taurine supplementation in experimental animal studies and habitual dietary patterns in humans. The absence of a GRS effect further suggests that genetically determined taurine metabolic capacity does not independently influence ageing trajectories in free-living human populations. This finding contrasts with mechanistic evidence from animal models. Intriguingly, while animal studies often report a decline in taurine levels with age, our data revealed that plasma taurine concentrations actually increased with age (data not shown). However, the consistent protective association of the plasma taurine/total AA ratio does not contradict the core biological premise proposed by Singh et al. [19] that taurine plays a functionally important role in ageing. Rather, it refines this concept by suggesting that ageing-related outcomes depend less on taurine intake or genetic capacity for synthesis and more on how efficiently it is maintained and regulated within the systemic amino acid milieu.

The paradoxical finding that higher dietary taurine-to-protein ratio was associated with increased PHAI-based unhealthy ageing risk, particularly in females (HR, 1.90), warrants careful mechanistic interpretation. The dissociation between dietary input and circulating taurine levels suggests that individuals consuming proportionally more dietary taurine may do so within a dietary pattern characterised by high seafood and meat intake, which simultaneously introduces other unfavourable components, such as advanced glycation end-products, saturated fatty acids, or excess methionine load through the transsulfuration pathway [36–39]. Furthermore, this dissociation is consistent with prior physiological evidence that dietary taurine contributes only modestly to systemic taurine status under habitual dietary conditions, given that endogenous biosynthesis via the transsulfuration pathway and renal reabsorption via *SLC6A6* are the principal determinants of circulating taurine homeostasis [10–12].

The sex-specific amplification of this paradoxical effect may reflect oestrogen-mediated suppression of hepatic *CDO1* and *CSAD* expression via oestrogen receptor-α, thereby reducing endogenous taurine biosynthetic capacity in females [40, 41]. Notably, this sex-specific effect persisted after adjustment for menopausal status, suggesting that the biological sex difference in taurine metabolic efficiency reflects fundamental, lifelong hormonal programming of taurine biosynthetic capacity rather than an acute consequence of oestrogen withdrawal. The adverse GRS × dietary taurine interaction in females further supports this interpretation. Among females with a high genetic taurine metabolic capacity, additional dietary taurine may paradoxically increase PHAI-based unhealthy ageing risk through amino acid metabolic imbalance or increased renal excretory burden.

The consistent protective association of the plasma taurine/total AA ratio, but not absolute taurine concentration, underscores the importance of evaluating taurine within the context of the entire amino acid pool. Absolute circulating taurine reflects the net balance of dietary intake, biosynthesis, and renal reabsorption, but is susceptible to confounding by overall nutritional status. In contrast, the taurine/total AA ratio reflects the relative abundance of taurine within the systemic amino acid pool, demonstrating the capacity of an organism to maintain taurine homeostasis even under conditions of metabolic demand. Individuals who maintain a high ratio may possess superior endogenous taurine retention through more efficient renal reabsorption via *SLC6A6*, greater biosynthetic efficiency via *CDO1* and *CSAD*, or more effective intracellular taurine cycling [5–7, 10–12]. This endogenous retention capacity may protect against multiple hallmarks of ageing, including mitochondrial dysfunction, oxidative stress, DNA damage, and inflammaging [19], by ensuring adequate taurine availability at the cellular level, even when absolute plasma concentrations are unremarkable.

The apparent paradox that participants in Q4 exhibited elevated waist circumference, blood pressure, and total cholesterol levels, yet showed significantly lower mortality and PHAI risk, than participants in the other quartiles is consistent with this resilience framework, [42, 43] and parallels the well-documented obesity paradox in older adults, wherein a mildly elevated BMI has been associated with lower mortality and more favourable ageing trajectories despite conventional cardiovascular risk profiles [44, 45].

The strengths of our study included its large prospective design with up to 14 years of follow-up and the unique simultaneous availability of plasma metabolomics, genome-wide genotyping, and repeated dietary assessment data within a single cohort. Methodologically, the use of the taurine/total AA ratio as a normalised index provided a more robust measure of taurine homeostasis than absolute plasma concentrations, accounting for overall nutritional status and amino acid pool size.

However, several limitations should be noted. First, metabolomics data were available for a subset of the KoGES cohort (n = 2,321), and plasma measurements were obtained at a single time point, precluding assessment of longitudinal changes in taurine homeostasis.

Second, dietary taurine intake was estimated via an FFQ, which may introduce measurement error and cannot capture short-term dietary variability.

Third, the study population was limited to community-dwelling ethnic Koreans, which may limit generalizability to other populations with different dietary patterns and genetic backgrounds.

Fourth, although dietary taurine intake was evaluated using a 1,000-kcal nutrient density approach, potential residual confounding from age- and sex-specific energy and protein requirements remains. Relatively younger individuals and males typically have higher requirements, which could affect nutrient density interpretability across subgroups. However, a standardised adjustment based on required density was limited due to the absence of established domestic or international dietary reference intakes or recommended nutrient intakes (RNIs) for taurine.

Fifth, although the transsulfuration pathway from methionine to taurine proceeds via cysteine and cysteine sulfinic acid intermediates catalysed by *CDO1* and *CSAD*, plasma cysteine and cysteine sulfinic acid concentrations were not available in the metabolomics panel, precluding direct assessment of upstream biosynthetic flux and limiting mechanistic inference regarding endogenous taurine production capacity.

Finally, although our findings provided robust evidence in a large human cohort, further longitudinal studies incorporating repeated metabolomics measurements, epigenetic profiling, and cross-population validation are required to fully characterise the translational potential of taurine homeostasis. Specifically, investigating DNA methylation patterns or histone modifications in taurine-related genes (e.g., *CDO1* and *CSAD*) could provide deeper insights into how environmental factors and ageing intersect with endogenous taurine regulation.

## Conclusions

The association between taurine and healthy ageing was not explained by dietary taurine intake or genetically determined taurine synthesis, but rather by its relative abundance within the systemic amino acid pool. These findings suggest that taurine functions as a biomarker of endogenous metabolic efficiency rather than as a deterministic causal driver. The sex-specific patterns observed (survival benefit among males and healthy ageing benefit among females) suggest that taurine homeostasis intersects with sex-specific ageing pathways, warranting further investigation into the underlying biological mechanisms.

Future longitudinal studies incorporating repeated metabolomics measurements, mechanistic investigation of renal taurine reabsorption efficiency, and intervention trials stratified by baseline taurine/total AA ratio are required to characterize the translational potential of taurine homeostasis as a target for healthy ageing.

## Data Availability

.KoGES data are available to qualified researchers through the
National Biobank of Korea (https://biobank.nih.go.kr) and the Clinical and Omics Data Archive (https://coda.nih.go.kr) upon approval of the research proposals and Institutional Review Board (IRB) clearance. Analytic code is available from the corresponding author upon reasonable request.

## Acknowledgements

We would like to thank all the participants and staff of the Korean Genome and Epidemiology Study (KoGES), without whom this study would not have been possible. This study was conducted with bioresources from National Biobank of Korea, The Korea Disease Control and Prevention Agency, Republic of Korea (NBK-2024-033).

## Author Contributions

All authors contributed to the conceptualization and methodology. Concept and design: Lyu, Lim, Park. Acquisition, analysis, or interpretation of data: Lyu, Park. Drafting of the manuscript: Lyu. Critical revision of the manuscript for important intellectual content: Lee, Hwang, Lim, Park. Statistical analysis: Lyu. Funding acquisition: Lim, Park. Supervision: Park.

## Data Sharing Statement

KoGES data are available to qualified researchers through the National Biobank of Korea (https://biobank.nih.go.kr) and the Clinical and Omics Data Archive (https://coda.nih.go.kr) upon approval of the research proposals and Institutional Review Board (IRB) clearance. Analytic code is available from the corresponding author upon reasonable request.

## Ethics Approval

The study was approved by the IRB (approval numbers: 2018-03-05-4C-A; 2022-07-07-C-A; ewha-202510-0042-01) and conducted in accordance with the Declaration of Helsinki. Written informed consent was obtained from all participants.

