## Supplementary Materials for "Plasma Taurine Relative Abundance, Not Dietary Intake or Genetic Predisposition, Predicts All-Cause Mortality and Unhealthy Ageing: A Prospective Cohort Study"

**Supplementary data**

Appendix 1. Sex-stratified hazard ratios for all three exposure variables for all-cause, age-related, cancer, and CVD mortality (Supplementary Tables S1–S3)

Appendix 2. Sensitivity analysis using plasma Taurine/Methionine ratio for mortality and PHAI-based unhealthy ageing (Supplementary Tables S4–S6)

Appendix 3. Sensitivity analyses using the extended 117-SNP taurine GRS incorporating CBS, CTH, and SLC36A2 genes (mortality and PHAI-based unhealthy ageing outcomes; total population and sex-stratified; Supplementary Tables S7–S9)

**Supplementary Tables S1. Sex-stratified multivariable-adjusted hazard ratios (HRs) and 95% confidence intervals (CIs) for all-cause and cause-specific mortality according to quartiles of plasma taurine-to-total amino acid ratio.**

|  | **Plasma Taurine/Total AA ratio** | | | | |
| --- | --- | --- | --- | --- | --- |
| **HRs (95% CIs)** | **Q1 (lowest)** | **Q2** | **Q3** | **Q4 (highest)** | **P for trend** |
| **Male** |  |  |  |  |  |
| No. of at risk | 282 | 282 | 283 | 282 |  |
| Person-years | 3,843 | 3,688 | 3,927 | 3,814 |  |
| Incidence rate per 1000 person-years | 13.01 | 13.29 | 10.19 | 12.06 |  |
| All–cause |  |  |  |  |  |
| No. of event | 50 | 49 | 40 | 46 |  |
| Crude model | Reference | 1.03 (0.69–1.52) | 0.76 (0.50–1.15) | 0.91 (0.61–1.36) | .4502 |
| Model 1 | Reference | 0.90 (0.61–1.34) | 0.67 (0.44–1.02) | 0.73 (0.49–1.09) | .0750 |
| Model 2 | Reference | 0.91 (0.61–1.36) | 0.65 (0.43–0.99) | 0.67 (0.44–1.00) | **.0273** |
| Age-related |  |  |  |  |  |
| No. of event | 44 | 39 | 37 | 41 |  |
| Crude model | Reference | 0.93 (0.61–1.43) | 0.81 (0.52–1.25) | 0.93 (0.61–1.42) | .6188 |
| Model 1 | Reference | 0.80 (0.52–1.24) | 0.71 (0.46–1.10) | 0.73 (0.47–1.11) | .1391 |
| Model 2 | Reference | 0.80 (0.52–1.24) | 0.68 (0.43–1.06) | 0.67 (0.43–1.03) | .0623 |
| Cancer |  |  |  |  |  |
| No. of event | 25 | 15 | 17 | 19 |  |
| Crude model | Reference | 0.63 (0.33–1.19) | 0.66 (0.36–1.22) | 0.76 (0.42–1.39) | .4060 |
| Model 1 | Reference | 0.55 (0.29–1.05) | 0.60 (0.32–1.11) | 0.61 (0.34–1.12) | .1475 |
| Model 2 | Reference | 0.55 (0.29–1.06) | 0.57 (0.31–1.07) | 0.58 (0.32–1.06) | .1004 |
| CVD |  |  |  |  |  |
| No. of event | 7 | 11 | 8 | 12 |  |
| Crude model | Reference | 1.65 (0.64–4.25) | 1.11 (0.40–3.06) | 1.74 (0.68–4.41) | .3503 |
| Model 1 | Reference | 1.41 (0.55–3.64) | 1.01 (0.37–2.79) | 1.35 (0.53–3.44) | .6670 |
| Model 2 | Reference | 1.27 (0.48–3.34) | 0.94 (0.34–2.64) | 1.15 (0.44–3.00) | .9085 |
| **Female** |  |  |  |  |  |
| No. of at risk | 298 | 298 | 298 | 298 |  |
| Person-years | 4,105 | 4,134 | 4,163 | 4,168 |  |
| Incidence rate per 1000 person-years | 6.33 | 7.74 | 8.41 | 8.64 |  |
| All–cause |  |  |  |  |  |
| No. of event | 26 | 32 | 35 | 36 |  |
| Crude model | Reference | 1.30 (0.77–2.19) | 1.26 (0.76–2.10) | 1.40 (0.84–2.31) | .2336 |
| Model 1 | Reference | 1.18 (0.70–1.98) | 1.10 (0.66–1.84) | 1.11 (0.67–1.84) | .7868 |
| Model 2 | Reference | 1.20 (0.70–2.04) | 1.07 (0.63–1.83) | 1.03 (0.61–1.73) | .9103 |
| Age-related |  |  |  |  |  |
| No. of event | 23 | 24 | 31 | 26 |  |
| Crude model | Reference | 1.13 (0.64–2.03) | 1.34 (0.77–2.31) | 1.19 (0.67–2.09) | .5923 |
| Model 1 | Reference | 0.99 (0.56–1.76) | 1.11 (0.65–1.92) | 0.89 (0.51–1.57) | .7325 |
| Model 2 | Reference | 1.02 (0.57–1.84) | 1.10 (0.62–1.94) | 0.82 (0.46–1.46) | .4781 |
| Cancer |  |  |  |  |  |
| No. of event | 12 | 10 | 10 | 10 |  |
| Crude model | Reference | 0.85 (0.37–1.98) | 0.81 (0.35–1.87) | 0.83 (0.36–1.92) | .6568 |
| Model 1 | Reference | 0.79 (0.34–1.83) | 0.73 (0.31–1.70) | 0.69 (0.30–1.60) | .3915 |
| Model 2 | Reference | 0.77 (0.32–1.82) | 0.75 (0.31–1.80) | 0.67 (0.28–1.61) | .3946 |
| CVD |  |  |  |  |  |
| No. of event | 8 | 5 | 10 | 10 |  |
| Crude model | Reference | 0.71 (0.23–2.18) | 1.11 (0.44–2.83) | 1.30 (0.51–3.31) | .4353 |
| Model 1 | Reference | 0.63 (0.21–1.93) | 0.88 (0.34–2.29) | 0.99 (0.39–2.51) | .8418 |
| Model 2 | Reference | 0.79 (0.24–2.58) | 0.81 (0.29–2.30) | 0.92 (0.34–2.51) | .9310 |

Q, Quartile; Q1: Quartile 1 (lowest), Q4: Quartile 4 (highest); CVD, cardiovascular disease; HR, hazard ratio; CI, confidence interval.

^a^ Crude model was unadjusted; Model 1 was adjusted for age (years, continuous); Model 2 was further adjusted for education level (elementary school or below, middle school, high school, or college or above), household income level (quartiles), smoking status (never, former, or current smoker), drinking status (never, former, or current drinker), regular physical activity (yes or no), body mass index (kg/m², continuous), total energy intake (kcal/day, continuous), and dietary taurine intake (mg/day, continuous). For the analysis in female, menopausal status (pre-menopausal vs. post-menopausal) was additionally adjusted in Model 2.

^b^ P for trend was calculated by assigning the median value of each quartile to participants as a continuous variable across all models. Bold values indicate statistical significance at P < 0.05.

**Supplementary Tables S2. Sex-stratified multivariable-adjusted hazard ratios (HRs) and 95% confidence intervals (CIs) for all-cause and cause-specific mortality according to quartiles of dietary taurine-to-protein ratio.**

|  | **Dietary Taurine/Protein ratio** | | | | |
| --- | --- | --- | --- | --- | --- |
| **HRs (95% CIs)** | **Q1 (lowest)** | **Q2** | **Q3** | **Q4 (highest)** | **P for trend** |
| **Male** |  |  |  |  |  |
| No. of at risk | 282 | 282 | 283 | 282 |  |
| Person-years | 3,771 | 3,774 | 3,906 | 3,821 |  |
| Incidence rate per 1000 person-years | 15.91 | 11.39 | 9.73 | 11.52 |  |
| All–cause |  |  |  |  |  |
| No. of event | 60 | 43 | 38 | 44 |  |
| Crude model | Reference | 0.75 (0.51–1.11) | 0.61 (0.41–0.92) | 0.73 (0.49–1.07) | .1068 |
| Model 1 | Reference | 1.00 (0.68–1.49) | 0.83 (0.55–1.25) | 0.90 (0.61–1.33) | .4870 |
| Model 2 | Reference | 1.09 (0.73–1.64) | 0.90 (0.59–1.38) | 0.92 (0.61–1.39) | .5832 |
| Age-related |  |  |  |  |  |
| No. of event | 55 | 39 | 31 | 36 |  |
| Crude model | Reference | 0.74 (0.49–1.11) | 0.54 (0.35–0.84) | 0.65 (0.43–0.99) | **.0347** |
| Model 1 | Reference | 1.00 (0.66–1.51) | 0.76 (0.49–1.18) | 0.83 (0.54–1.27) | .2798 |
| Model 2 | Reference | 1.13 (0.74–1.73) | 0.83 (0.52–1.31) | 0.86 (0.55–1.34) | .3665 |
| Cancer |  |  |  |  |  |
| No. of event | 28 | 21 | 17 | 10 |  |
| Crude model | Reference | 0.76 (0.43–1.34) | 0.58 (0.32–1.07) | 0.35 (0.17–0.73) | **.0029** |
| Model 1 | Reference | 1.00 (0.57–1.77) | 0.79 (0.43–1.45) | 0.45 (0.22–0.94) | **.0253** |
| Model 2 | Reference | 1.12 (0.62–2.02) | 0.88 (0.47–1.67) | 0.48 (0.23–1.02) | **.0467** |
| CVD |  |  |  |  |  |
| No. of event | 8 | 10 | 6 | 14 |  |
| Crude model | Reference | 1.26 (0.50–3.18) | 0.72 (0.25–2.08) | 1.73 (0.73–4.12) | .2319 |
| Model 1 | Reference | 1.77 (0.70–4.48) | 1.06 (0.37–3.08) | 2.38 (1.00–5.69) | .0733 |
| Model 2 | Reference | 2.40 (0.89–6.47) | 1.20 (0.40–3.63) | 2.50 (0.99–6.34) | .1033 |
| **Female** |  |  |  |  |  |
| No. of at risk | 298 | 298 | 298 | 298 |  |
| Person-years | 4,243 | 4,147 | 4,076 | 4,104 |  |
| Incidence rate per 1000 person-years | 8.25 | 10.13 | 5.89 | 6.82 |  |
| All–cause |  |  |  |  |  |
| No. of event | 35 | 42 | 24 | 28 |  |
| Crude model | Reference | 1.15 (0.73–1.81) | 0.65 (0.38–1.09) | 0.78 (0.47–1.28) | .1094 |
| Model 1 | Reference | 1.35 (0.85–2.12) | 1.01 (0.60–1.71) | 1.19 (0.72–1.96) | .8016 |
| Model 2 | Reference | 1.35 (0.85–2.15) | 1.08 (0.63–1.85) | 1.22 (0.72–2.07) | .6989 |
| Age-related |  |  |  |  |  |
| No. of event | 32 | 33 | 17 | 22 |  |
| Crude model | Reference | 0.99 (0.61–1.62) | 0.50 (0.28–0.91) | 0.67 (0.39–1.16) | .0530 |
| Model 1 | Reference | 1.17 (0.71–1.92) | 0.81 (0.45–1.46) | 1.05 (0.61–1.81) | .8525 |
| Model 2 | Reference | 1.13 (0.68–1.88) | 0.85 (0.46–1.56) | 1.00 (0.56–1.79) | .7963 |
| Cancer |  |  |  |  |  |
| No. of event | 9 | 15 | 5 | 13 |  |
| Crude model | Reference | 1.66 (0.72–3.79) | 0.56 (0.19–1.66) | 1.45 (0.62–3.41) | .7862 |
| Model 1 | Reference | 1.93 (0.84–4.43) | 0.78 (0.26–2.36) | 2.02 (0.86–4.76) | .2803 |
| Model 2 | Reference | 1.83 (0.79–4.25) | 0.74 (0.24–2.28) | 1.85 (0.75–4.54) | .4041 |
| CVD |  |  |  |  |  |
| No. of event | 10 | 10 | 6 | 7 |  |
| Crude model | Reference | 0.87 (0.36–2.12) | 0.49 (0.17–1.38) | 0.61 (0.23–1.63) | .2470 |
| Model 1 | Reference | 0.96 (0.38–2.40) | 0.88 (0.31–2.46) | 1.07 (0.40–2.83) | .9141 |
| Model 2 | Reference | 0.96 (0.36–2.53) | 0.93 (0.31–2.73) | 1.02 (0.35–2.94) | .9718 |

Q, Quartile; Q1: Quartile 1 (lowest), Q4: Quartile 4 (highest); CVD, cardiovascular disease; HR, hazard ratio; CI, confidence interval.

^a^ Crude model was unadjusted; Model 1 was adjusted for age (years, continuous); Model 2 was further adjusted for education level (elementary school or below, middle school, high school, or college or above), household income level (quartiles), smoking status (never, former, or current smoker), drinking status (never, former, or current drinker), regular physical activity (yes or no), body mass index (kg/m², continuous), and total energy intake (kcal/day, continuous). For the analysis in female, menopausal status (pre-menopausal vs. post-menopausal) was additionally adjusted in Model 2.

^b^ P for trend was calculated by assigning the median value of each quartile to participants as a continuous variable across all models. Bold values indicate statistical significance at P < 0.05.

**Supplementary Tables S3. Sex-stratified multivariable-adjusted hazard ratios (HRs) and 95% confidence intervals (CIs) for all-cause and cause-specific mortality according to quartiles of the genetic risk score (GRS) for taurine metabolism and transport.**

|  | **Genetic Risk Score (GRS)** | | | | |
| --- | --- | --- | --- | --- | --- |
| **HRs (95% CIs)** | **Q1 (lowest)** | **Q2** | **Q3** | **Q4 (highest)** | **P for trend** |
| **Male** |  |  |  |  |  |
| No. of at risk | 264 | 263 | 264 | 263 |  |
| Person-years | 3,637 | 3,684 | 3,768 | 3,651 |  |
| Incidence rate per 1000 person-years | 11.82 | 10.59 | 10.88 | 14.24 |  |
| All–cause |  |  |  |  |  |
| No. of event | 43 | 39 | 41 | 52 |  |
| Crude model | Reference | 0.84 (0.54–1.29) | 0.88 (0.57–1.35) | 1.18 (0.79–1.77) | .4121 |
| Model 1 | Reference | 0.79 (0.51–1.23) | 0.79 (0.52–1.22) | 1.06 (0.70–1.59) | .7904 |
| Model 2 | Reference | 0.86 (0.55–1.34) | 0.88 (0.57–1.36) | 1.13 (0.75–1.71) | .5560 |
| Age-related |  |  |  |  |  |
| No. of event | 37 | 33 | 36 | 47 |  |
| Crude model | Reference | 0.84 (0.52–1.34) | 0.91 (0.57–1.44) | 1.25 (0.81–1.92) | .2895 |
| Model 1 | Reference | 0.80 (0.50–1.28) | 0.83 (0.52–1.31) | 1.12 (0.72–1.72) | .5983 |
| Model 2 | Reference | 0.87 (0.54–1.41) | 0.90 (0.57–1.44) | 1.20 (0.77–1.87) | .4151 |
| Cancer |  |  |  |  |  |
| No. of event | 15 | 16 | 14 | 26 |  |
| Crude model | Reference | 1.04 (0.51–2.10) | 0.89 (0.43–1.85) | 1.72 (0.91–3.25) | .1100 |
| Model 1 | Reference | 0.99 (0.49–1.99) | 0.83 (0.40–1.72) | 1.58 (0.84–2.98) | .1809 |
| Model 2 | Reference | 1.02 (0.50–2.08) | 0.89 (0.43–1.86) | 1.63 (0.85–3.13) | .1548 |
| CVD |  |  |  |  |  |
| No. of event | 13 | 7 | 8 | 10 |  |
| Crude model | Reference | 0.53 (0.21–1.34) | 0.59 (0.25–1.43) | 0.77 (0.34–1.76) | .4884 |
| Model 1 | Reference | 0.52 (0.21–1.30) | 0.55 (0.23–1.34) | 0.70 (0.31–1.59) | .3572 |
| Model 2 | Reference | 0.66 (0.26–1.70) | 0.69 (0.28–1.71) | 0.82 (0.35–1.95) | .6075 |
| **Female** |  |  |  |  |  |
| No. of at risk | 272 | 273 | 269 | 276 |  |
| Person-years | 3,972 | 3,975 | 3,837 | 3,867 |  |
| Incidence rate per 1000 person-years | 7.05 | 8.05 | 9.90 | 6.72 |  |
| All–cause |  |  |  |  |  |
| No. of event | 28 | 32 | 38 | 26 |  |
| Crude model | Reference | 1.08 (0.65–1.80) | 1.48 (0.91–2.41) | 1.04 (0.61–1.78) | .5418 |
| Model 1 | Reference | 1.02 (0.61–1.70) | 1.26 (0.77–2.05) | 1.05 (0.62–1.80) | .6355 |
| Model 2 | Reference | 0.97 (0.58–1.63) | 1.21 (0.74–1.99) | 1.05 (0.61–1.82) | .6301 |
| Age-related |  |  |  |  |  |
| No. of event | 22 | 24 | 31 | 23 |  |
| Crude model | Reference | 1.00 (0.56–1.80) | 1.54 (0.89–2.66) | 1.17 (0.65–2.10) | .3494 |
| Model 1 | Reference | 0.97 (0.54–1.73) | 1.28 (0.74–2.22) | 1.18 (0.65–2.11) | .4112 |
| Model 2 | Reference | 0.89 (0.49–1.61) | 1.23 (0.71–2.14) | 1.19 (0.65–2.16) | .3934 |
| Cancer |  |  |  |  |  |
| No. of event | 10 | 8 | 10 | 12 |  |
| Crude model | Reference | 0.77 (0.31–1.97) | 1.06 (0.44–2.54) | 1.28 (0.55–2.98) | .4853 |
| Model 1 | Reference | 0.76 (0.30–1.92) | 0.94 (0.39–2.26) | 1.28 (0.55–2.97) | .5283 |
| Model 2 | Reference | 0.78 (0.30–2.01) | 0.92 (0.38–2.24) | 1.42 (0.60–3.37) | .4343 |
| CVD |  |  |  |  |  |
| No. of event | 9 | 11 | 9 | 3 |  |
| Crude model | Reference | 1.10 (0.45–2.67) | 1.13 (0.45–2.85) | 0.40 (0.11–1.49) | .2998 |
| Model 1 | Reference | 0.98 (0.40–2.38) | 0.88 (0.35–2.23) | 0.40 (0.11–1.49) | .2245 |
| Model 2 | Reference | 0.75 (0.29–1.97) | 0.83 (0.32–2.18) | 0.35 (0.09–1.37) | .1847 |

Q, quartile (Q1, lowest; Q4, highest); HR, hazard ratio; CI, confidence interval; CVD, cardiovascular disease; GRS, genetic risk score.

^a^ Crude model was unadjusted; Model 1 was adjusted for age (years, continuous); Model 2 was further adjusted for education level (elementary school or below, middle school, high school, or college or above), household income level (quartiles), smoking status (never, former, or current smoker), drinking status (never, former, or current drinker), regular physical activity (yes or no), body mass index (kg/m², continuous), total energy intake (kcal/day, continuous), and protein intake(g/day, continuous). For the analysis in female, menopausal status (pre-menopausal vs. post-menopausal) was additionally adjusted in Model 2.

^b^ P for trend was calculated by assigning the median value of each quartile of the GRS to participants as a continuous variable across all models.

**Supplementary Tables S4. Multivariable-adjusted hazard ratios (HRs) and 95% confidence intervals (CIs) for all-cause and cause-specific mortality according to quartiles of plasma taurine-to-methionine ratio.**

|  | **Plasma Taurine/Methionine ratio** | | | | |
| --- | --- | --- | --- | --- | --- |
| **HRs (95% CIs)** | **Q1 (lowest)** | **Q2** | **Q3** | **Q4 (highest)** | **P for trend** |
| No. of at risk | 580 | 580 | 581 | 580 |  |
| Person-years | 7,863 | 7,974 | 8,124 | 7,881 |  |
| Incidence rate per 1000 person-years | 11.19 | 7.90 | 9.60 | 10.79 |  |
| All–cause |  |  |  |  |  |
| No. of event | 88 | 63 | 78 | 85 |  |
| Crude model | Reference | 0.74 (0.53–1.02) | 0.87 (0.64–1.19) | 0.98 (0.73–1.32) | .7478 |
| Model 1 | Reference | 0.65 (0.47–0.90) | 0.73 (0.53–1.00) | 0.80 (0.59–1.10) | .4485 |
| Model 2 | Reference | 0.64 (0.46–0.89) | 0.68 (0.50–0.93) | 0.72 (0.53–0.98) | .1325 |
| Age-related |  |  |  |  |  |
| No. of event | 77 | 53 | 67 | 68 |  |
| Crude model | Reference | 0.70 (0.49–1.00) | 0.85 (0.61–1.18) | 0.89 (0.64–1.24) | .8198 |
| Model 1 | Reference | 0.61 (0.43–0.87) | 0.71 (0.51–0.99) | 0.73 (0.52–1.02) | .2092 |
| Model 2 | Reference | 0.61 (0.43–0.87) | 0.66 (0.47–0.93) | 0.66 (0.46–0.93) | .0596 |
| Cancer |  |  |  |  |  |
| No. of event | 39 | 22 | 26 | 31 |  |
| Crude model | Reference | 0.56 (0.33–0.95) | 0.65 (0.39–1.06) | 0.80 (0.50–1.28) | .5487 |
| Model 1 | Reference | 0.52 (0.31–0.89) | 0.59 (0.36–0.98) | 0.71 (0.44–1.17) | .3575 |
| Model 2 | Reference | 0.52 (0.31–0.89) | 0.54 (0.32–0.89) | 0.63 (0.38–1.04) | .1471 |
| CVD |  |  |  |  |  |
| No. of event | 17 | 12 | 20 | 22 |  |
| Crude model | Reference | 0.73 (0.35–1.53) | 1.16 (0.61–2.22) | 1.31 (0.70–2.47) | .2079 |
| Model 1 | Reference | 0.59 (0.28–1.25) | 0.88 (0.46–1.71) | 0.96 (0.50–1.84) | .6862 |
| Model 2 | Reference | 0.59 (0.28–1.25) | 0.88 (0.45–1.73) | 0.87 (0.44–1.71) | .9231 |

Q, quartile (Q1, lowest; Q4, highest); HR, hazard ratio; CI, confidence interval; CVD, cardiovascular disease.

^a^ Crude model was unadjusted; Model 1 was adjusted for age (years, continuous) and sex; Model 2 was further adjusted for education level (elementary school or below, middle school, high school, or college or above), household income level (quartiles), smoking status (never, former, or current smoker), drinking status (never, former, or current drinker), regular physical activity (yes or no), body mass index (kg/m², continuous), total energy intake (kcal/day, continuous), and dietary taurine intake (mg/day, continuous).

^b^ P for trend was calculated by assigning the median value of each quartile to participants as a continuous variable across all models.

**Supplementary Tables S5. Sex-stratified multivariable-adjusted hazard ratios (HRs) and 95% confidence intervals (CIs) for all-cause and cause-specific mortality according to quartiles of plasma taurine-to-methionine ratio.**

| **Plasma Taurine/Methionine ratio** | | | | | |
| --- | --- | --- | --- | --- | --- |
| **HRs (95% CIs)** | **Q1 (lowest)** | **Q2** | **Q3** | **Q4 (highest)** | **P for trend** |
| **Male** |  |  |  |  |  |
| No. of at risk | 282 | 282 | 283 | 282 |  |
| Person-years | 3,906 | 3,693 | 3,889 | 3,784 |  |
| Incidence rate per 1000 person-years | 12.80 | 12.73 | 8.74 | 14.27 |  |
| All–cause |  |  |  |  |  |
| No. of event | 50 | 47 | 34 | 54 |  |
| Crude model | Reference | 1.05 (0.71–1.57) | 0.70 (0.45–1.08) | 1.10 (0.75–1.62) | .8388 |
| Model 1 | Reference | 0.96 (0.64–1.43) | 0.53 (0.34–0.82) | 0.80 (0.54–1.18) | .1395 |
| Model 2 | Reference | 0.94 (0.62–1.41) | 0.47 (0.30–0.73) | 0.68 (0.46–1.02) | **.0238** |
| Age-related |  |  |  |  |  |
| No. of event | 43 | 42 | 28 | 48 |  |
| Crude model | Reference | 1.09 (0.71–1.67) | 0.66 (0.41–1.07) | 1.15 (0.76–1.74) | .7370 |
| Model 1 | Reference | 0.98 (0.64–1.50) | 0.49 (0.30–0.79) | 0.81 (0.53–1.22) | .1657 |
| Model 2 | Reference | 0.96 (0.62–1.48) | 0.44 (0.27–0.71) | 0.70 (0.46–1.08) | **.0449** |
| Cancer |  |  |  |  |  |
| No. of event | 20 | 20 | 12 | 24 |  |
| Crude model | Reference | 1.08 (0.58–2.01) | 0.61 (0.30–1.24) | 1.25 (0.69–2.26) | .6203 |
| Model 1 | Reference | 0.99 (0.53–1.84) | 0.46 (0.22–0.94) | 0.90 (0.49–1.63) | .5499 |
| Model 2 | Reference | 0.94 (0.50–1.77) | 0.38 (0.18–0.80) | 0.77 (0.42–1.42) | .2860 |
| CVD |  |  |  |  |  |
| No. of event | 9 | 7 | 9 | 11 |  |
| Crude model | Reference | 0.83 (0.31–2.24) | 1.00 (0.40–2.53) | 1.51 (0.65–3.53) | .2382 |
| Model 1 | Reference | 0.75 (0.28–2.01) | 0.72 (0.29–1.82) | 1.02 (0.44–2.41) | .7897 |
| Model 2 | Reference | 0.67 (0.24–1.86) | 0.67 (0.26–1.78) | 0.91 (0.37–2.24) | .9682 |
| **Female** |  |  |  |  |  |
| No. of at risk | 298 | 298 | 298 | 298 |  |
| Person-years | 4,094 | 4,248 | 4,177 | 4,051 |  |
| Incidence rate per 1000 person-years | 8.30 | 4.00 | 9.82 | 9.13 |  |
| All–cause |  |  |  |  |  |
| No. of event | 34 | 17 | 41 | 37 |  |
| Crude model | Reference | 0.49 (0.28–0.88) | 1.19 (0.76–1.88) | 1.11 (0.70–1.77) | .1514 |
| Model 1 | Reference | 0.40 (0.22–0.72) | 0.97 (0.62–1.53) | 0.85 (0.54–1.36) | .5864 |
| Model 2 | Reference | 0.42 (0.23–0.76) | 0.95 (0.59–1.52) | 0.83 (0.52–1.24) | .7790 |
| Age-related |  |  |  |  |  |
| No. of event | 30 | 10 | 36 | 28 |  |
| Crude model | Reference | 0.34 (0.17–0.70) | 1.22 (0.75–2.00) | 0.99 (0.59–1.67) | .3278 |
| Model 1 | Reference | 0.26 (0.13–0.54) | 0.96 (0.59–1.56) | 0.72 (0.43–1.20) | .9086 |
| Model 2 | Reference | 0.28 (0.14–0.58) | 0.93 (0.56–1.54) | 0.69 (0.41–1.17) | .8618 |
| Cancer |  |  |  |  |  |
| No. of event | 13 | 5 | 15 | 9 |  |
| Crude model | Reference | 0.38 (0.13–1.05) | 1.13 (0.54–2.38) | 0.70 (0.30–1.64) | .8786 |
| Model 1 | Reference | 0.33 (0.12–0.93) | 0.99 (0.47–2.08) | 0.57 (0.24–1.33) | .5457 |
| Model 2 | Reference | 0.30 (0.10–0.85) | 0.89 (0.42–1.91) | 0.53 (0.22–1.25) | .4480 |
| CVD |  |  |  |  |  |
| No. of event | 12 | 1 | 9 | 11 |  |
| Crude model | Reference | 0.09 (0.01–0.66) | 0.74 (0.31–1.76) | 0.94 (0.41–2.13) | .5038 |
| Model 1 | Reference | 0.06 (0.01–0.44) | 0.53 (0.22–1.28) | 0.69 (0.31–1.57) | .7896 |
| Model 2 | Reference | 0.07 (0.01–0.54) | 0.49 (0.19–1.25) | 0.66 (0.28–1.57) | .9914 |

Q, Quartile; Q1: Quartile 1 (lowest), Q4: Quartile 4 (highest); CVD, cardiovascular disease; HR, hazard ratio; CI, confidence interval.

^a^ Crude model was unadjusted; Model 1 was adjusted for age (years, continuous); Model 2 was further adjusted for education level (elementary school or below, middle school, high school, or college or above), household income level (quartiles), smoking status (never, former, or current smoker), drinking status (never, former, or current drinker), regular physical activity (yes or no), body mass index (kg/m², continuous), total energy intake (kcal/day, continuous), and dietary taurine intake (mg/day, continuous). For the analysis in female, menopausal status (pre-menopausal vs. post-menopausal) was additionally adjusted in Model 2.

^b^ P for trend was calculated by assigning the median value of each quartile to participants as a continuous variable across all models. Bold values indicate statistical significance at P < 0.05.

**Supplementary Tables S6. Multivariable-adjusted hazard ratios (HRs) and 95% confidence intervals (CIs) for the incidence of PHAI-based unhealthy ageing according to quartiles of plasma taurine-to-methionine ratio.**

|  | **Plasma Taurine/Methionine ratio** | | | | |
| --- | --- | --- | --- | --- | --- |
| **HRs (95% CIs)** | **Q1 (lowest)** | **Q2** | **Q3** | **Q4 (highest)** | **P for trend** |
| **All** |  |  |  |  |  |
| No. of at risk | 296 | 296 | 296 | 296 |  |
| No. of event | 121 | 132 | 123 | 119 |  |
| Person-years | 2,669 | 2,637 | 2,651 | 2,719 |  |
| Incidence rate per 1000 person-years | 45.34 | 50.05 | 46.40 | 43.76 |  |
| Crude model | Reference | 1.11 (0.87–1.43) | 1.02 (0.79–1.31) | 0.96 (0.75–1.24) | .5597 |
| Model 1 | Reference | 1.12 (0.87–1.44) | 0.95 (0.74–1.23) | 0.93 (0.71–1.21) | .3388 |
| Model 2 | Reference | 1.03 (0.80–1.33) | 0.85 (0.66–1.11) | 0.81 (0.62–1.06) | .0575 |
| **Male** |  |  |  |  |  |
| No. of at risk | 137 | 137 | 137 | 137 |  |
| No. of event | 57 | 63 | 62 | 62 |  |
| Person-years | 1 216 | 1 170 | 1 176 | 1 191 |  |
| Incidence rate per 1000 person-years | 46.89 | 53.87 | 52.73 | 52.08 |  |
| Crude model | Reference | 1.18 (0.82–1.70) | 1.14 (0.79–1.65) | 1.13 (0.78–1.62) | .6251 |
| Model 1 | Reference | 1.25 (0.87–1.80) | 1.09 (0.75–1.56) | 1.07 (0.74–1.53) | .9956 |
| Model 2 | Reference | 1.13 (0.78–1.65) | 0.89 (0.61–1.30) | 0.95 (0.64–1.40) | .5158 |
| **Female** |  |  |  |  |  |
| No. of at risk | 159 | 159 | 160 | 159 |  |
| No. of event | 65 | 60 | 63 | 63 |  |
| Person-years | 1 486 | 1 518 | 1 444 | 1 477 |  |
| Incidence rate per 1000 person-years | 43.74 | 39.53 | 43.64 | 42.66 |  |
| Crude model | Reference | 0.90 (0.64–1.28) | 0.97 (0.69–1.38) | 0.95 (0.67–1.34) | .8961 |
| Model 1 | Reference | 0.90 (0.63–1.28) | 0.90 (0.64–1.28) | 0.83 (0.59–1.17) | .3283 |
| Model 2 | Reference | 0.82 (0.58–1.17) | 0.80 (0.56–1.14) | 0.69 (0.48–1.00) | .0611 |

Q, quartile (Q1, lowest; Q4, highest); HR, hazard ratio; CI, confidence interval; PHAI, Physiology Healthy Ageing Index.

^a^ Crude model was unadjusted; Model 1 was adjusted for age (years, continuous), with sex additionally adjusted in the analysis of the total population; Model 2 was further adjusted for education level (elementary school or below, middle school, high school, or college or above), household income level (quartiles), smoking status (never, former, or current smoker), drinking status (never, former, or current drinker), regular physical activity (yes or no), body mass index (kg/m², continuous), total energy intake (kcal/day, continuous), and dietary taurine intake (mg/day, continuous). For the analysis in female, menopausal status (pre-menopausal vs. post-menopausal) was additionally adjusted in Model 2.

^b^ P for trend was calculated by assigning the median value of each quartile of plasma taurine-to-total amino acid ratio to participants as a continuous variable across all models.

^c^ Unhealthy ageing was defined as a PHAI score at or below the 25th percentile (≤25th) of the study population at follow-up. Participants who met the criteria for unhealthy ageing at baseline were excluded from this analysis.

**Supplementary Tables S7. Multivariable-adjusted hazard ratios (HRs) and 95% confidence intervals (CIs) for all-cause and cause-specific mortality according to quartiles of the expanded 117-SNP taurine GRS.**

|  | **Taurine Genetic Risk Score (GRS).** | | | | |
| --- | --- | --- | --- | --- | --- |
| **HRs (95% CIs)** | **Q1 (lowest)** | **Q2** | **Q3** | **Q4 (highest)** | **P for trend** |
| No. of at risk | 424 | 424 | 424 | 424 |  |
| Person-years | 5,818 | 5,935 | 5,706 | 5,705 |  |
| Incidence rate per 1000 person-years | 10.14 | 9.44 | 11.92 | 10.69 |  |
| All–cause |  |  |  |  |  |
| No. of event | 59 | 56 | 68 | 61 |  |
| Crude model | Reference | 0.91 (0.63–1.31) | 1.15 (0.81–1.62) | 1.07 (0.75–1.53) | .5051 |
| Model 1 | Reference | 0.85 (0.59–1.22) | 1.09 (0.77–1.55) | 0.99 (0.69–1.41) | .7889 |
| Model 2 | Reference | 0.84 (0.58–1.22) | 1.09 (0.77–1.55) | 1.01 (0.70–1.45) | .7112 |
| Age-related |  |  |  |  |  |
| No. of event | 42 | 53 | 55 | 52 |  |
| Crude model | Reference | 1.21 (0.81–1.82) | 1.31 (0.87–1.95) | 1.28 (0.85–1.91) | .1989 |
| Model 1 | Reference | 1.11 (0.74–1.67) | 1.22 (0.82–1.83) | 1.17 (0.78–1.76) | .3683 |
| Model 2 | Reference | 1.09 (0.72–1.64) | 1.21 (0.81–1.82) | 1.18 (0.78–1.78) | .3535 |
| Cancer |  |  |  |  |  |
| No. of event | 19 | 22 | 24 | 24 |  |
| Crude model | Reference | 1.12 (0.61–2.07) | 1.27 (0.70–2.32) | 1.29 (0.71–2.36) | .3542 |
| Model 1 | Reference | 1.04 (0.56–1.93) | 1.20 (0.66–2.19) | 1.22 (0.67–2.22) | .4589 |
| Model 2 | Reference | 1.08 (0.58–2.01) | 1.22 (0.66–2.23) | 1.25 (0.68–2.30) | .4204 |
| CVD |  |  |  |  |  |
| No. of event | 25 | 14 | 18 | 14 |  |
| Crude model | Reference | 1.15 (0.58–2.29) | 1.06 (0.53–2.15) | 0.76 (0.35–1.65) | .5579 |
| Model 1 | Reference | 1.05 (0.53–2.08) | 1.00 (0.49–2.02) | 0.69 (0.32–1.50) | .4009 |
| Model 2 | Reference | 0.99 (0.49–2.00) | 1.01 (0.49–2.07) | 0.77 (0.35–1.68) | .5798 |

Q, quartile (Q1, lowest; Q4, highest); HR, hazard ratio; CI, confidence interval; CVD, cardiovascular disease; GRS, genetic risk score.

^a^ Crude model was unadjusted; Model 1 was adjusted for age (years, continuous) and sex; Model 2 was further adjusted for education level (elementary school or below, middle school, high school, or college or above), household income level (quartiles), smoking status (never, former, or current smoker), drinking status (never, former, or current drinker), regular physical activity (yes or no), body mass index (kg/m², continuous), total energy intake (kcal/day, continuous), and protein intake(g/day, continuous).

^b^ P for trend was calculated by assigning the median value of each quartile of the GRS to participants as a continuous variable across all models.

**Supplementary Tables S8. Sex-stratified multivariable-adjusted hazard ratios (HRs) and 95% confidence intervals (CIs) for all-cause and cause-specific mortality according to quartiles of the expanded 117-SNP taurine GRS.**

|  | **Taurine Genetic Risk Score (GRS).** | | | | |
| --- | --- | --- | --- | --- | --- |
| **HRs (95% CIs)** | **Q1 (lowest)** | **Q2** | **Q3** | **Q4 (highest)** | **P for trend** |
| **Male** |  |  |  |  |  |
| No. of at risk | 213 | 213 | 214 | 213 |  |
| Person-years | 2,820 | 2,991 | 2,802 | 2,828 |  |
| Incidence rate per 1000 person-years | 12.77 | 11.03 | 15.35 | 12.02 |  |
| All–cause |  |  |  |  |  |
| No. of event | 36 | 33 | 43 | 34 |  |
| Crude model | Reference | 0.87 (0.54–1.40) | 1.19(0.76–1.85) | 0.95 (0.59–1.52) | .8631 |
| Model 1 | Reference | 0.83 (0.52–1.34) | 1.04 (0.67–1.62) | 0.90 (0.56–1.44) | .8626 |
| Model 2 | Reference | 0.77 (0.47–1.26) | 1.00 (0.64–1.57) | 0.91 (0.56–1.46) | .8919 |
| Age-related |  |  |  |  |  |
| No. of event | 26 | 32 | 36 | 30 |  |
| Crude model | Reference | 1.16 (0.69–1.95) | 1.37 (0.83–2.27) | 1.15 (0.68–1.94) | .4571 |
| Model 1 | Reference | 1.10 (0.65–1.84) | 1.18 (0.71–1.96) | 1.09 (0.64–1.84) | .6775 |
| Model 2 | Reference | 1.02 (0.60–1.74) | 1.15 (0.69–1.92) | 1.06 (0.62–1.82) | .7068 |
| Cancer |  |  |  |  |  |
| No. of event | 14 | 15 | 17 | 13 |  |
| Crude model | Reference | 1.00 (0.48–2.07) | 1.21 (0.59–2.45) | 0.91 (0.43–1.95) | .9766 |
| Model 1 | Reference | 0.94 (0.46–1.96) | 1.04 (0.51–2.11) | 0.87 (0.41–1.86) | .8075 |
| Model 2 | Reference | 0.95 (0.45–1.99) | 1.06 (0.52–2.17) | 0.87 (0.40–1.87) | .8157 |
| CVD |  |  |  |  |  |
| No. of event | 6 | 7 | 10 | 7 |  |
| Crude model | Reference | 1.09 (0.37–3.23) | 1.66 (0.61–4.58) | 1.15 (0.39–3.42) | .6046 |
| Model 1 | Reference | 1.03 (0.35–3.06) | 1.40 (0.51–3.86) | 1.11 (0.37–3.30) | .7046 |
| Model 2 | Reference | 0.68 (0.21–2.15) | 1.09 (0.38–3.11) | 1.09 (0.36–3.30) | .7144 |
| **Female** |  |  |  |  |  |
| No. of at risk | 210 | 211 | 211 | 211 |  |
| Person-years | 2,975 | 2,952 | 2,911 | 2,885 |  |
| Incidence rate per 1000 person-years | 8.07 | 7.45 | 8.59 | 9.36 |  |
| All–cause |  |  |  |  |  |
| No. of event | 24 | 22 | 25 | 27 |  |
| Crude model | Reference | 0.85 (0.48–1.53) | 1.02 (0.58–1.78) | 1.18 (0.68–2.05) | .5313 |
| Model 1 | Reference | 0.81 (0.45–1.44) | 1.11 (0.63–1.94) | 1.05 (0.61–1.83) | .6751 |
| Model 2 | Reference | 0.78 (0.43–1.42) | 1.11 (0.62–2.00) | 0.98 (0.56–1.71) | .8653 |
| Age-related |  |  |  |  |  |
| No. of event | 17 | 20 | 19 | 22 |  |
| Crude model | Reference | 1.11 (0.58–2.12) | 1.10 (0.57–2.11) | 1.36 (0.72–2.55) | .3843 |
| Model 1 | Reference | 1.04 (0.54–1.99) | 1.19 (0.62–2.30) | 1.20 (0.64–2.27) | .5086 |
| Model 2 | Reference | 0.97 (0.50–1.90) | 1.22 (0.61–2.42) | 1.15 (0.60–2.19) | .5790 |
| Cancer |  |  |  |  |  |
| No. of event | 4 | 8 | 7 | 11 |  |
| Crude model | Reference | 1.94 (0.58–6.46) | 1.75 (0.51–5.98) | 2.85 (0.91–8.96) | .0845 |
| Model 1 | Reference | 1.88 (0.57–6.25) | 1.86 (0.54–6.37) | 2.72 (0.86–8.54) | .0915 |
| Model 2 | Reference | 1.92 (0.57–6.46) | 1.62 (0.46–5.69) | 2.74 (0.86–8.72) | .1060 |
| CVD |  |  |  |  |  |
| No. of event | 9 | 11 | 6 | 4 |  |
| Crude model | Reference | 1.08 (0.44–2.63) | 0.63 (0.22–1.78) | 0.47 (0.14–1.52) | .1598 |
| Model 1 | Reference | 0.99 (0.41–2.41) | 0.71 (0.25–2.00) | 0.37 (0.11–1.23) | .1011 |
| Model 2 | Reference | 1.06 (0.41–2.73) | 0.87 (0.28–2.67) | 0.42 (0.12–1.41) | .1929 |

Q, quartile (Q1, lowest; Q4, highest); HR, hazard ratio; CI, confidence interval; CVD, cardiovascular disease; GRS, genetic risk score.

^a^ Crude model was unadjusted; Model 1 was adjusted for age (years, continuous); Model 2 was further adjusted for education level (elementary school or below, middle school, high school, or college or above), household income level (quartiles), smoking status (never, former, or current smoker), drinking status (never, former, or current drinker), regular physical activity (yes or no), body mass index (kg/m², continuous), total energy intake (kcal/day, continuous), and protein intake(g/day, continuous). For the analysis in female, menopausal status (pre-menopausal vs. post-menopausal) was additionally adjusted in Model 2.

^b^ P for trend was calculated by assigning the median value of each quartile of the GRS to participants as a continuous variable across all models.

**Supplementary Tables S9. Multivariable-adjusted hazard ratios (HRs) and 95% confidence intervals (CIs) for the incidence of PHAI-based unhealthy ageing according to quartiles of the expanded 117-SNP taurine GRS.**

|  | **Taurine Genetic Risk Score (GRS).** | | | | |
| --- | --- | --- | --- | --- | --- |
| **HRs (95% CIs)** | **Q1 (lowest)** | **Q2** | **Q3** | **Q4 (highest)** | **P for trend** |
| **All** |  |  |  |  |  |
| No. of at risk | 296 | 296 | 296 | 296 |  |
| No. of event | 121 | 132 | 123 | 119 |  |
| Person-years | 2,669 | 2,637 | 2,651 | 2,719 |  |
| Incidence rate per 1000 person-years | 45.34 | 50.05 | 46.40 | 43.76 |  |
| Crude model | Reference | 1.11 (0.87–1.43) | 1.02 (0.79–1.31) | 0.96 (0.75–1.24) | .5597 |
| Model 1 | Reference | 1.12 (0.87–1.44) | 0.95 (0.74–1.23) | 0.93 (0.71–1.21) | .3388 |
| Model 2 | Reference | 1.03 (0.80–1.33) | 0.85 (0.66–1.11) | 0.81 (0.62–1.06) | .0575 |
| **Male** |  |  |  |  |  |
| No. of at risk | 137 | 137 | 137 | 137 |  |
| No. of event | 57 | 63 | 62 | 62 |  |
| Person-years | 1 216 | 1 170 | 1 176 | 1 191 |  |
| Incidence rate per 1000 person-years | 46.89 | 53.87 | 52.73 | 52.08 |  |
| Crude model | Reference | 1.18 (0.82–1.70) | 1.14 (0.79–1.65) | 1.13 (0.78–1.62) | .6251 |
| Model 1 | Reference | 1.25 (0.87–1.80) | 1.09 (0.75–1.56) | 1.07 (0.74–1.53) | .9956 |
| Model 2 | Reference | 1.13 (0.78–1.65) | 0.89 (0.61–1.30) | 0.95 (0.64–1.40) | .5158 |
| **Female** |  |  |  |  |  |
| No. of at risk | 159 | 159 | 160 | 159 |  |
| No. of event | 65 | 60 | 63 | 63 |  |
| Person-years | 1,486 | 1,518 | 1,444 | 1,477 |  |
| Incidence rate per 1000 person-years | 43.74 | 39.53 | 43.64 | 42.66 |  |
| Crude model | Reference | 0.90 (0.64–1.28) | 0.97 (0.69–1.38) | 0.95 (0.67–1.34) | .8961 |
| Model 1 | Reference | 0.90 (0.63–1.28) | 0.90 (0.64–1.28) | 0.83 (0.59–1.17) | .3283 |
| Model 2 | Reference | 0.82 (0.58–1.17) | 0.80 (0.56–1.14) | 0.69 (0.48–1.00) | .0611 |

Q, quartile (Q1, lowest; Q4, highest); HR, hazard ratio; CI, confidence interval; PHAI, Physiology Healthy Ageing Index.

^a^ Crude model was unadjusted; Model 1 was adjusted for age (years, continuous), with sex additionally adjusted in the analysis of the total population; Model 2 was further adjusted for education level (elementary school or below, middle school, high school, or college or above), household income level (quartiles), smoking status (never, former, or current smoker), drinking status (never, former, or current drinker), regular physical activity (yes or no), body mass index (kg/m², continuous), total energy intake (kcal/day, continuous), and protein intake(g/day, continuous). For the analysis in female, menopausal status (pre-menopausal vs. post-menopausal) was additionally adjusted in Model 2. ^b^ P for trend was calculated by assigning the median value of each quartile of plasma taurine-to-total amino acid ratio to participants as a continuous variable across all models. ^c^ Unhealthy ageing was defined as a PHAI score at or below the 25th percentile (≤25th) of the study population at follow-up. Participants who met the criteria for unhealthy ageing at baseline were excluded from this analysis.
